# High Burden of Shigellosis Among Hospitalized Children under five in Ndola, Zambia: Evidence from molecular-based surveillance

**DOI:** 10.64898/2026.10.29.26366813

**Authors:** Sam Miti, Seter Siziya, Roma Chilengi, Catherine Zulu, Naomi Muleba Kalaba, Christian Musilikare Niyongabo, Chipo Manda, Lydia Chisapi, Moses Chakopo, Roy Moono, Mwizukanji Nachamba, Innocent Chilumba, Bwendo Nduna, Lusungu Msimuko, David Nick Thole, Gershom Chongwe, Lungowe Mukubuta, Aliness Mwilaba Dombola, Muleya Inambao, Goren Sofya, Imasiku Ilubala, Mwelwa Chibuye Michelo Simuyandi, Caroline Cleopatra Chisenga, Dani Cohen

**Affiliations:** Copperbelt University, Michael Chilufya School of Medicine, Ndola, Zambia; National Health Research and Training Institute (formerly Tropical Diseases Research Centre), Ndola, Zambia; Arthur Davison Children’s Hospital, Ndola, Zambia; Center for Infectious Disease Research in Zambia (CIDRZ), Lusaka, Zambia; Zambia National Public Health Institute, Lusaka, Zambia; School of Public Health, Gray Faculty of Medical & Health Sciences, Tel Aviv University, Tel Aviv, Israel

**Keywords:** *Shigella*, Shigellosis, Bacillary Dysentery, Diarrhea, Children, Epidemiology, Incidence, Prevalence, Loop-mediated isothermal amplification (LAMP), Zambia, Sub-Saharan Africa

## Abstract

**Background:** Shigella spp. is a leading cause of moderate-to-severe diarrhea and diarrheal mortality in children under five (CU5), globally. Traditional culture-based techniques consistently underestimates its prevalence, particularly in high-burden regions of sub-Saharan Africa. We aimed to generate population-relevant estimates of shigellosis among hospitalized children under five (CU5) in Ndola, Zambia, using a sensitive molecular diagnostic approach.

**Methods:** A prospective, hospital-based surveillance study was conducted at Arthur Davison Children’s Hospital (ADCH) from May 2020 to November 2021. All CU5 admitted with acute diarrhea (≥2 watery stools/day) were eligible. From enrolled children, sociodemographic and clinical data were collected, and stool samples were obtained where possible. Primary detection of *Shigella* spp./enteroinvasive *E. coli* (EIEC) was performed using loop-mediated isothermal amplification (LAMP) targeting the conserved *ipaH* gene. Operational shigellosis case-definition: A child with diarrhea (2 or more watery stools/day) and a stool sample positive for the ipaH gene by LAMP. Prevalence of shigellosis was calculated among children with LAMP-tested samples and weighted to account for all admissions. The incidence of hospitalization was estimated using the district CU5 population denominators.

**Results:** Among 1,685 enrolled children, 950 provided stool samples. LAMP detected *ipaH* in 186 samples (19.6%;95% CI: 17.1–22.5). The weighted prevalence among all admissions was 19.5% (95% CI: 17.5–21.5) corresponding to 329 cases. The incidence of shigellosis hospitalization was 2.4 (95% CI: 2.2–2.7) per 1,000 person-years. The burden exhibited a distinct age distribution, peaking in children aged 12–23 months, preceded by an evident rise in the number of cases between 6 and 12 months of age. Shigellosis was associated with bloody diarrhea, increased stool frequency, and fever. Pre-admission antibiotic or other medication use was reported in 33% of cases.

**Conclusions:** Shigellosis accounts for a substantial proportion of severe diarrhea requiring hospitalization in Ndola, Zambia. Findings support improved diagnostics, antimicrobial stewardship, and prioritization of Shigella vaccines.

## Introduction

Diarrheal diseases persist as a leading killer of children under five (CU5) globally, with an estimated 444,000 deaths in 2021, the vast majority occurring in low- and middle-income countries (LMICs) of sub-Saharan Africa and South Asia (1–3). While the etiological landscape of diarrhea is diverse, encompassing viral, bacterial, and parasitic pathogens, *Shigella* spp. stand out due to their low infectious dose, propensity to cause severe inflammatory dysentery, and significant contribution to mortality and long-term sequelae such as stunting (2,4,5). The Global Burden of Disease Study 2023 estimated that diarrhoeal diseases caused 1.11 million (95% UI 0.811–1.54) deaths worldwide in 2023, of which *Shigella* spp. accounted for 9.3% (5.4–15.2), making it the leading bacterial cause of diarrhoeal mortality, behind only rotavirus and norovirus overall. Among children younger than 5 years, *Shigella* was responsible for 24.0% (15.1–36.7) of diarrhoeal deaths (5–7).

Historically, quantifying the burden of shigellosis has been hampered by diagnostic limitations. Conventional culture, the longstanding gold standard, suffers from poor sensitivity. This is exacerbated in real-world settings where antibiotic usage prior to seeking care is common, further reducing bacterial viability (8,9). The landmark Global Enteric Multicenter Study (GEMS) and subsequent reanalysis using quantitative polymerase chain reaction (qPCR) unveiled a critical truth: culture-based methods significantly underestimate the burden of *Shigella*, as well as other enteric pathogens (10–12). The qPCR reanalysis of GEMS samples nearly doubled the attributable fraction of moderate-to-severe diarrhea for *Shigella*, demonstrating that a substantial proportion of disease is missed by traditional microbiology (12,13). Additionally, Miti et al found that utilization of clinical dysentery as a proxy of shigellosis grossly under-estimated the burden of shigellosis in Zambian children (13). This diagnostic gap obscures the true scale of the problem, potentially leading to its under-prioritization in public health agendas and vaccine development roadmaps (9).

In Zambia, as in many African nations, data on shigellosis are sparse and historically linked to outbreaks, such as the multi-drug resistant *Shigella dysenteriae* type 1 epidemic in the 1990s (14,15). Recent studies employing molecular diagnostics in Lusaka, Zambia’s capital, have identified *Shigella* as a major contributor to childhood diarrhea (16). However, a comprehensive understanding of its hospital-based burden, age-specific incidence, and clinical profile outside the capital is lacking. Such granular, context-specific data is needed. It informs local treatment guidelines, shapes rational antibiotic policy, identifies high-risk groups for targeted interventions, and provides the foundational epidemiology necessary for planning and powering future vaccine trials (17–20).

Ndola, a major urban center on the Copperbelt, serves as a relevant sentinel site. Arthur Davison Children’s Hospital (ADCH) functions as the primary pediatric referral center for the district. Conducting surveillance here captures cases of severe diarrhea representative of the broader community. This study aimed to leverage a sensitive molecular tool—loop-mediated isothermal amplification (LAMP)—to overcome the limitations of culture and generate estimates of the prevalence, population-based incidence, and epidemiological characteristics of shigellosis among CU5 admitted with diarrhea in Ndola, Zambia. The findings are intended to bridge a critical knowledge gap by presenting new data alongside its limitations, offering national and global health stakeholders balanced considerations for action against Shigella associated diarrhea.

## Methods

### Study Design and Setting

A prospective, hospital-based surveillance study was conducted at Arthur Davison Children’s Hospital (ADCH) in Ndola, Zambia, from May 1, 2020, to November 30, 2021. ADCH is a government-run tertiary facility that served as the sole dedicated pediatric referral hospital for Ndola district, effectively functioning as the primary admission center for severe childhood illness in its catchment area at the time of the study. The study period encompassed phases of the COVID-19 pandemic, but ADCH remained the central point for severe pediatric care.

### Study Population and Enrollment

The target population was children aged 0-59 months residing in Ndola district who presented to ADCH and were admitted with a primary diagnosis of acute diarrhea, defined as the passage of two or more watery stools in a 24-hour period. Children with known chronic gastrointestinal conditions or those admitted for non-diarrheal illnesses were excluded. The estimated mid-year population of CU5 in Ndola district during the surveillance period, extrapolated from the 2017 census projection of 86,097, was used for incidence calculations. A complete enumeration approach was employed in which all eligible children admitted during the surveillance period were included in the surveillance to minimize selection bias.

### Data and Specimen Collection

Demographic and clinical data were routinely obtained as part of routine diarrhea surveillance data by clinicians and nurses using a routine hospital diarrhea surveillance form. The surveillance data captured detailed sociodemographic information (age, sex, residence), clinical history (duration of illness, stool frequency, presence of blood, fever, vomiting, pre-admission medication use), and physical examination findings (hydration status, axillary temperature, weight). Data were supplemented by abstraction from the child’s medical file. A fresh stool sample was collected from the child whenever possible. For children who could not provide a sample at admission, rectal swabs were not used; thus, sample collection was contingent on natural defecation only. Stool samples were immediately tested using loop-mediated isothermal amplification (LAMP) and an aliquot stored at -80°C .

### Laboratory Testing – Primary Diagnostic

The primary diagnostic method was a LAMP assay targeting the invasion plasmid antigen H (*ipaH*) gene, which is present in all *Shigella* species and enteroinvasive *Escherichia coli* (EIEC) (21,22). The LAMP assay was performed according to a previously validated protocol (22,23). This method was chosen for its high sensitivity, specificity, and operational advantages (isothermal reaction, visual readout) suitable for resource-limited settings. All testing was performed in a centralized laboratory.

### Case definition

A child with diarrhea (2 or more watery stools/day) and a stool sample positive for the ipaH gene by LAMP was defined as a case of shigellosis. This was the operational case-definition used based on prior speciation and metagenomics findings indicating that EIEC—which also harbors the 220 kb virulence plasmid and the ipaH gene—is substantially less common than *Shigella* (10).

### Statistical Analysis

Data were entered into a DHIS2 database and analyzed using IBM SPSS software version 29 (IBM, Armonk US). Descriptive statistics summarized the characteristics of the enrolled cohort. Prevalence of shigellosis was calculated as the proportion of children with diarrhea and LAMP-ipaH positive results among the total of children with diarrhea and stool samples tested for ipaH by LAMP. To estimate the prevalence of shigellosis among all admitted CU5 (n=1,685) and account for missing stool samples tested by LAMP, a weighted prevalence was calculated. Weights were assigned as the inverse of the monthly sampling fraction (number tested/number enrolled), applied separately for each month of the study period. Incidence of hospitalization was calculated by dividing the weighted number of estimated cases (329) by the person-time at risk. Person-time was estimated as the product of the district CU5 population (86,097) and the surveillance period (1.58 years), yielding incidence per 1,000 person-years with approximate 95% confidence intervals (CI) derived from the Poisson distribution. Associations between shigellosis status and categorical variables (e.g., age group, sex, fever) were assessed by logistic regression with statistical significance testing using the chi-square or Fisher’s exact tests. For the antibiotic treatment variable, we conducted SAS Firth’s bias correction with odds ratios (OR) and Profile-Likelihood (PL) confidence Intervals. A p-value of <0.05 was considered statistically significant

### Ethical Considerations

The study protocol was reviewed and approved by the National Health Research Authority (24) and the Tropical Diseases Research Ethics Committee (25). All data were anonymized prior to analysis.

## Results

### Characteristics of the Study Population

Over the 19-month surveillance period, 1,685 CU5 admitted with acute diarrhea were surveyed by the study. Stool samples were successfully obtained and tested by LAMP from 950 children (56.4% sampling fraction). (Fig 1).

**Fig 1.**
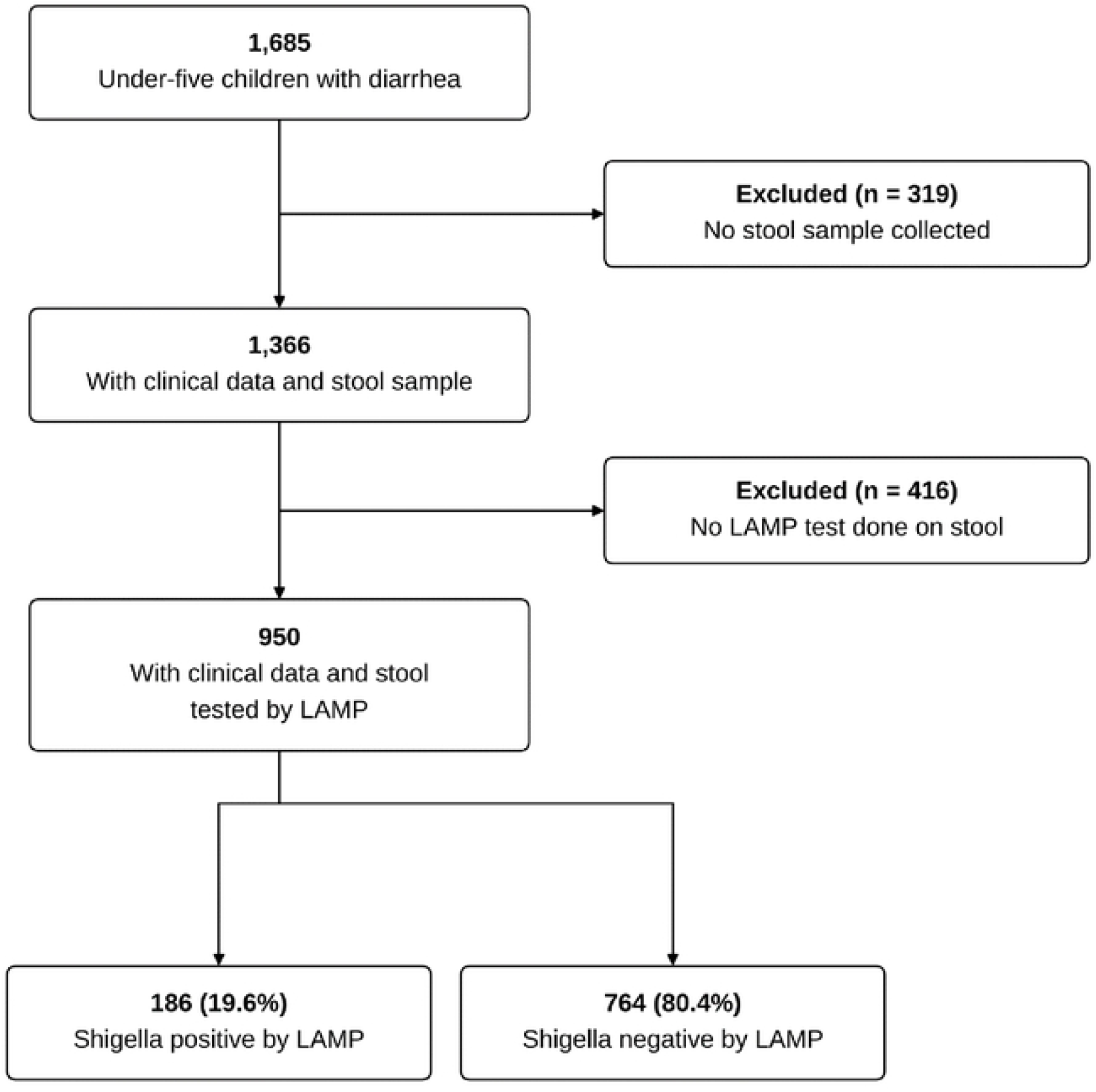
Flowchart of diarrhea case inclusion and Shigella testing results. Legend Figure 1: Flowchart showing stool sample availability, and testing for Shigella among children admitted with diarrhoea in Ndola, Zambia, from May 2020 to November 2021. Of 950 children whose stool samples were tested, 186 (19.6%) were positive by loop-mediated isothermal amplification (LAMP).

The sociodemographic profile of children who provided a stool sample was comparable to that of the entire enrolled cohort, suggesting the tested sample was representative (Table 1). The median age was 16 months (IQR: 10-24), and 55% of the children were male.

**Table 1:** Socio-demographic characteristics of all enrolled diarrhea cases and the subset with stool tested by LAMP.

|  | **LAMP tested for *Shigella***<br>(N=950) |  | **Total admitted with diarrhea**<br>(N=1685) |  |
|---|---|---|---|---|
| **Variable** | **N** | **%** | **n** | **%** |
| **Sex** |  |  |  |  |
| Male | 539 | 56.7 | 963 | 57.2 |
| Female | 411 | 43.3 | 722 | 42.8 |
| **Age** |  |  |  |  |
| 0-12 | 439 | 46.2 | 743 | 44.1 |
| 13-24 | 306 | 32.2 | 555 | 32.9 |
| 25-59 | 205 | 21.6 | 387 | 23.0 |
| **Residence** |  |  |  |  |
| High density, low economic status | 611 | 64.3 | 1080 | 64.1 |
| Medium/low density, high-medium economic status | 269 | 28.3 | 471 | 28.0 |
| Rural setting/Agricultural setting/Other | 70 | 7.4 | 134 | 8.0 |

### Prevalence of shigellosis

The LAMP assay detected the *ipaH* gene in 186 of the 950tested stooled stool samples, resulting in an unadjusted prevalence of 19.6% (95% CI: 17.1–22.5). After applying inverse probability weighting to account for children without a stool sample, the estimated prevalence of shigellosis among all 1,685 hospitalized diarrhea cases was 19.5% 95% CI: 17.5–21.5). This corresponds to an estimated 329 children admitted with shigellosis during the study period.

### Epidemiological Correlates

The prevalence of shigellosis exhibited a significant variation by age (p<0.001) as illustrated in Table 2., When the five-year under-five age range was considered by year of life, the greatest concentration of shigellosis cases occurred during the second year of life (12–23 months). This was preceded by a marked increase in the cumulative number of cases beginning between 6 and 12 months of age (Figure 2), indicating that the burden rises substantially during late infancy and is greatest, in terms of annual case count, during the second year of life.”

**Table 2:** Prevalence overall and demographic characteristics of shigellosis.

| **Variable** | **Total with diarrhea** | **ipaH negative n (%)** | **ipaH positive n (%)** | **OR (95% CI)** | **P-value** |
|---|---|---|---|---|---|
| **All** | 950 | 764 (80.4) | n=186 (19.6) |  |  |
| **Sex** |  |  |  |  |  |
| Male | 539 | 440 (57.6) | 99 (53.2) | 0.84 (0.61-1.16) | 0.28 |
| Female | 411 | 324 (42.4) | 87 (46.8) | Reference |  |
| **Age** |  |  |  |  |  |
| 0-12 months | 439 | 389 (50.9) | 50 (26.9) | Reference |  |
| 13-24 months | 306 | 238 (31.2) | 68 (36.6) | 2.22 (1.49-3.31) | **<0.001** |
| 25-59 months | 205 | 137 (17.9) | 68 (36.6) | 3.86 (2.55-5.84) | **<0.001** |
| **Residence** |  |  |  |  |  |
| High density, low economic status | 611 | 484 (63.4) | 127 (68.3) | Reference |  |
| Medium/low density, high-medium economic status | 269 | 223 (29.2) | 46 (24.7) | 0.79 (0.54-1.14) | 0.21 |
| Rural setting/Agricultural setting/Other | 70 | 57 (7.5) | 13 (7.0) | 0.87 (0.46-1.64) | 0.66 |
| **Antibiotic treatment at admission** |  |  |  |  |  |
| Didn't get antibiotics | 21 | 20 (2.7) | 1 (0.5) | Reference |  |
| Got recommended antibiotics | 162 | 133 (18.0) | 29 (15.9) | 3.02 (0.72-27.93) | 0.21 |
| Got antibiotics not recommended for AGE (penicillin X) | 737 | 585 (79.3) | 152 (83.5) | 3.56 (0.90-32.22) | 0.15 |

Missing data: antibiotic treatment (n=30); Percentages are calculated excluding missing values.
Prevalence out of tested: 186/950=19.6% (95% CI=17.1-22.5).  There was no significant difference in prevalence between males and females (p=0.28).  However age was a significant factor (P <0.001).

**Fig 2.**
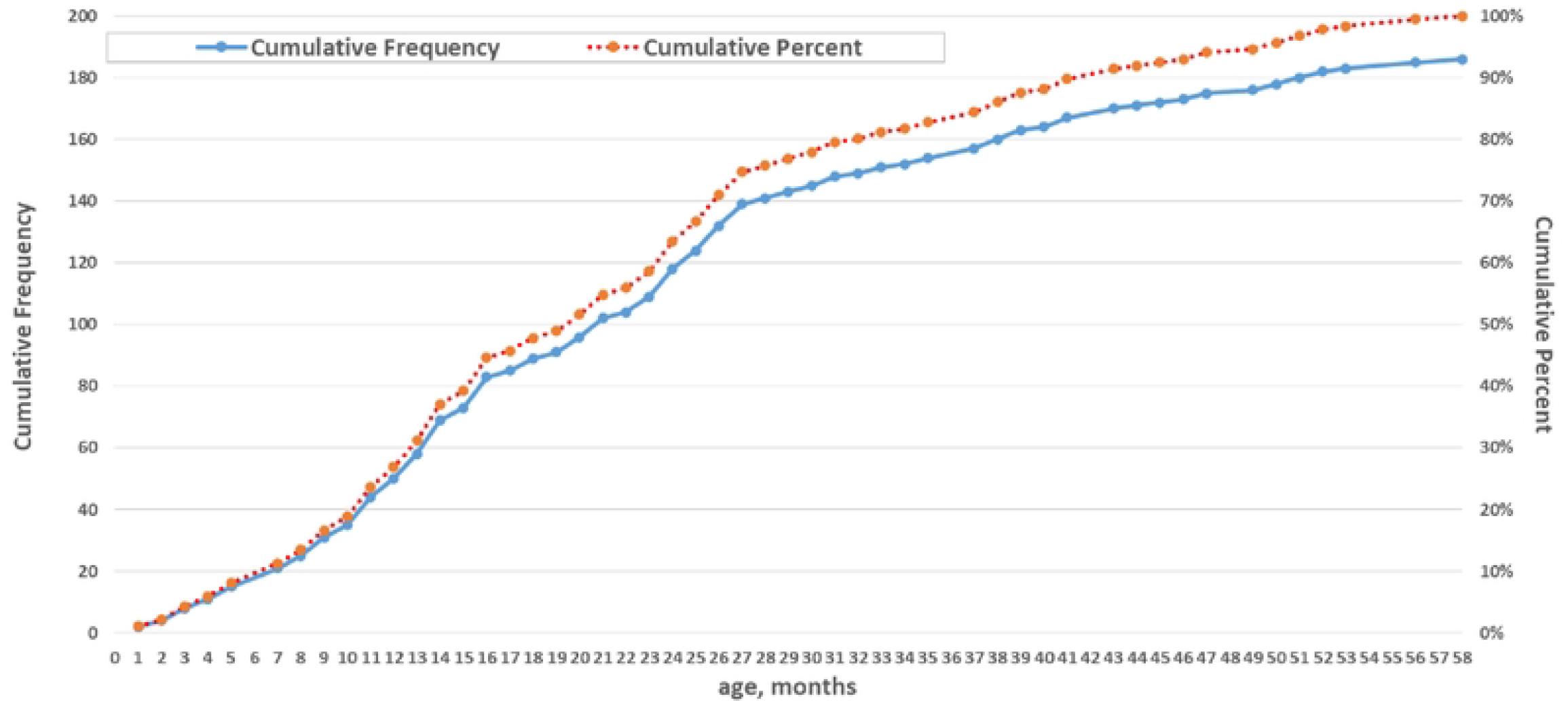
Cumulative distribution by age of the 186 children with LAMP defined shigellosis. Legend Figure 2: Cumulative age distribution of the 186 children with LAMP-defined shigellosis. The solid blue line shows the cumulative number of cases (left axis), and the dotted orange line shows the cumulative percentage (right axis). Each point represents the number or percentage of cases at or below the corresponding age in months.

### Clinical Presentation and Management

Clinical characteristics of shigellosis are summarized in Table 3. Using watery diarrhea as the reference category, children presenting with bloody diarrhea were over four times more likely to have shigellosis (OR 4.21, 95% CI 2.27–7.79), while mucoid diarrhea was also associated with increased odds of shigellosis (OR 1.56, 95% CI 1.07–2.26). Compared with the ipaH-negative cases of diarrhea, children with shigellosis were significantly more likely to present with bloody diarrhea (11.3% vs 3.5%; OR 4.21, 95% CI 2.27–7.79), while watery diarrhea was less common and used as the reference category (p < 0.001). Shigellosis was also associated with a higher number of stools per day, with increased odds among children with 3–7 episodes/day (OR 2.13, 95% CI 1.22–3.71) and ≥8 episodes/day (OR 2.60, 95% CI 1.24–5.49) compared with <3 episodes/day (p = 0.014). A history of vomiting was less frequent among shigellosis cases (OR 0.69, 95% CI 0.49–0.98; p = 0.048), whereas a history of fever at admission was more common (OR 1.77, 95% CI 1.18– 2.66; p = 0.005). A significant finding was that 33% of caregivers reported administering antibiotics or other medications before seeking hospital care.

**Table 3:** Clinical characteristics of children with shigellosis.

| **Variable** | **Total with diarrhea** | **ipaH negative (%)** | **ipaH positive (%)** | **OR** | **95% CI** | **P-value** |
|---|---|---|---|---|---|---|
| **Type of diarrhoea** | (n=950) | (n=764) | (n=186) |  |  |  |
| Watery | 590 | 495 (65.8) | 95 (51.1) | Reference |  |  |
| Bloody | 47 | 26 (3.5) | 21 (11.3) | 4.21 | 2.27-7.79 | **<0.001** |
| Mucoid | 239 | 184 (24.5) | 55 (29.6) | 1.56 | 1.07-2.26 | **0.020** |
| rice water | 29 | 21 (2.8) | 8 (4.3) | 1.99 | 0.85-4.61 | 0.111 |
| Other | 33 | 26 (3.5) | 7 (3.8) | 1.40 | 0.59-3.33 | 0.442 |
| **Number of diarrheic stools per day** |  |  |  |  |  |  |
| < 3 episodes/day | 141 | 125 (19.4) | 16 (9.9) | Reference |  |  |
| 3-7 episodes/day | 593 | 466 (72.2) | 127 (78.9) | 2.13 | 1.22-3.71 | **0.008** |
| 8 or more episodes/day | 72 | 54 (8.4) | 18 (11.2) | 2.60 | 1.24-5.49 | **0.012** |
| **History of vomiting at admission** |  |  |  |  |  |  |
| No | 240 | 182 (23.8) | 58 (31.2) | Reference |  |  |
| Yes | 710 | 582 (76.2) | 128 (68.8) | 0.69 | 0.49-0.98 | **0.039** |
| **History of fever at admission** |  |  |  |  |  |  |
| No | 251 | 217 (28.4) | 34 (18.3) | Reference |  |  |
| Yes | 699 | 547 (71.6) | 152 (81.7) | 1.77 | 1.18-2.66 | **0.005** |

Missing data: type of diarrhea (n=12), number of diarrheic stools per day (n=144),
Percentages are calculated excluding missing values.

### Temporal Trends

The monthly positivity rate for *Shigella* fluctuated between 10% and 30% throughout the surveillance period (Fig 3 and 4). A seasonal pattern was discernible during the 19-month observation window, with peaks during the hot, dry seasons.

**Fig 3.**
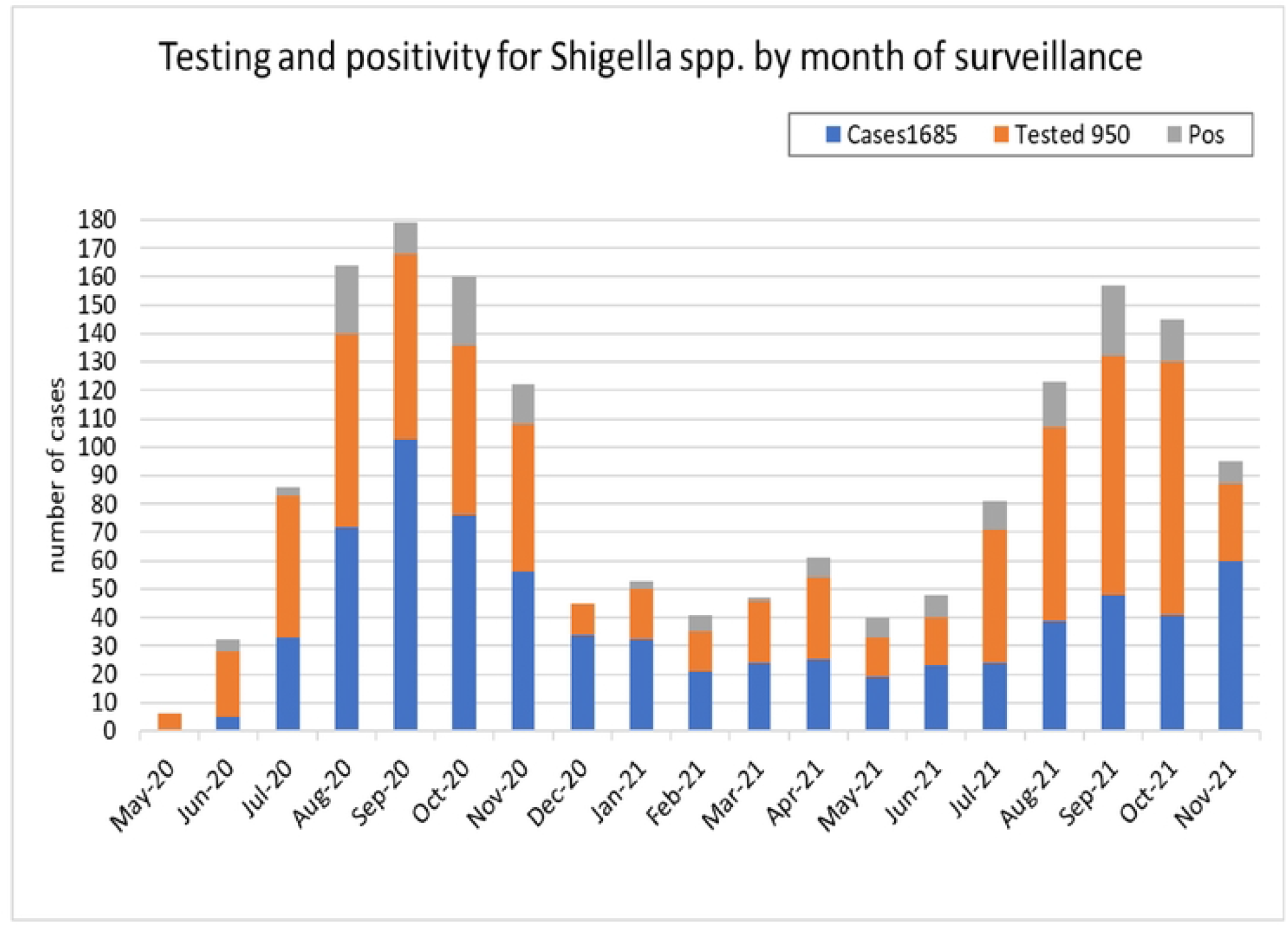
Shigellosis by month of surveillance (May 2020 – Nov 2021) Legend Fig 3: Weighted prevalence. 329/1685=19.5% (95%CI=17.5-21.5) Weighted using the inverse of the sampling fraction with weights attributed separately to each month of admission

**Fig 4.**
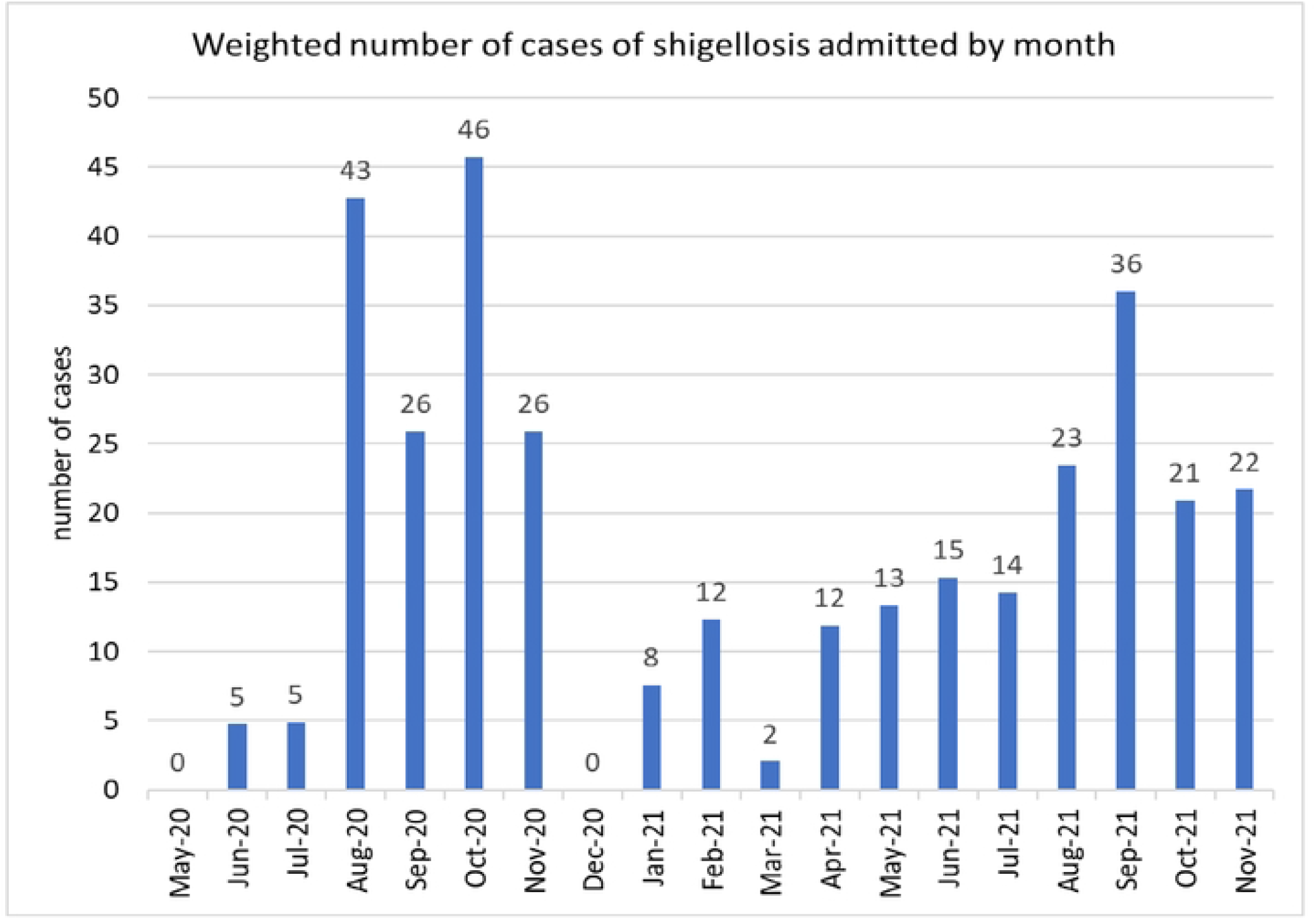
Estimated Incidence of shigellosis associated with hospitalization. Legend Fig 4: Estimated monthly numbers of shigellosis admissions after weighting, from May 2020 to November 2021. Bars represent weighted case estimates, and labels show the corresponding values rounded to the nearest whole number.

Using the weighted case estimate of 329 and a denominator of 136,320 person-years (86,097 children * 1.58 years), the incidence of hospitalization for LAMP-proven shigellosis was calculated to be 2.4 (95% CI: 2.2–2.7) per 1,000 person-years among CU5 in Ndola district.

## Discussion

This hospital-based surveillance study, employing a sensitive molecular assay, reveals a \staggering burden of shigellosis among CU5 in Ndola, Zambia. A weighted prevalence of 19.5% indicates that approximately one in five hospitalizations for childhood diarrhea met the operational molecular case definition for shigellosis.” This figure is substantially higher than many historical culture-based or clinical dysentery reports from Zambia(13,16) and sub-Saharan Africa and aligns with the revised, higher estimates from the qPCR reanalysis of the GEMS study, which found *Shigella* to be grossly under-estimated and a top pathogen in African sites(12,26). Our findings confirm that in high-transmission settings, *Shigella* is not a minor contributor but a leading etiology of severe pediatric diarrhea, demanding commensurate public health attention.

The estimated incidence of 2.4 hospitalizations per 1,000 person-years provides an important population-level metric. It translates to a substantial absolute number of severe cases requiring intensive healthcare resources in a district with a finite CU5 population. This incidence is comparable to or exceeds rates reported for other vaccine-preventable severe childhood infections in the region, underscoring the potential impact of an effective *Shigella* vaccine (27,28). The concentration of cases in the second year of life (12-24 months) is a consistent and critical epidemiological feature observed in LMICs globally(29,30), and it was preceded by an evident rise in the number of cases between 6 and 12 months of age with a cumulative frequency of 26.9% at 12 months.

Interestingly, this is in line with the extent of exposure to *Shigella* detected by serology in a cohort of Zambian infants in Lusaka (31). This window represents a period of heightened vulnerability due to weaning (increased exposure to contaminated food and water), developmental mobility (crawling, handling objects), and the waning of protective maternal antibodies. This precise age targeting is important for designing vaccination schedules, suggesting that primary immunization with a *Shigella* vaccine should ideally be completed as early as possible toward the end of the first year of life to provide protection during this window and in the subsequent years of risk (32,33).

A substantial proportion of participants (33%) reported antibiotic or other medication use prior to admission. This practice has multiple important implications. First, it illustrates why culture-based surveillance fails to show high prevalence of shigellosis, as antibiotics drastically reduce bacterial viability and thus detection (37,38). Our use of

LAMP, which detects pathogen DNA regardless of viability, was essential to circumvent this bias and providing a more sensitive estimate of the disease burden. Second, it reflects widespread inappropriate antibiotic use for acute watery diarrhea, which is most often viral or, in the case of *Shigella*, often not indicated for mild cases. This fuels the global crisis of antimicrobial resistance (AMR), which is particularly relevant for *Shigella* (39,40). Third, early antibiotic clearance may blunt the natural immune response to Shigella, with potential implications for the development of natural immunity.

Clinical associations observed in this study, including bloody diarrhea, fever, and increased stool frequency, are consistent with the invasive nature of Shigella infection. However, the majority of ipaH-positive cases did not present with dysentery, supporting evidence that reliance on clinical dysentery alone underestimates the burden of Shigella-associated disease (13).

The clinical severity observed among *Shigella*-positive cases—high fever, frequent stools, dehydration—reinforces its role as a pathogen of public health importance that strains clinical services. While our study design does not calculate a population-based mortality rate, the severity of hospitalized cases suggests that *Shigella* likely contributes to diarrheal mortality in this setting, consistent with global burden estimates (5,6).

Our study has limitations. As a hospital-based study, it captures the burden of severe disease but not the likely larger burden of mild or moderate community cases. The *ipaH* gene target does not differentiate between *Shigella* species and EIEC, nor does it provide serotype information directly. However we based our operational shigellosis case definition on previous evidence indicating that EIEC is substantially less common than *Shigella* in similar settings. Finally, the sampling fraction of 56.4%, while addressed by weighting, represents a potential source of bias, though the demographic comparability of tested and untested children is reassuring. The increased shigellosis prevalence during the hot/dry months in the 19-month observation period is notable but still insufficient to establish a reproducible seasonal pattern.

## Conclusion

Shigellosis represents a substantial burden among children under five requiring hospitalization in Ndola, Zambia. The high prevalence, age-specific risk, and association with severe clinical features highlight its public health importance. Further studies to determine the specific *Shigella* serotypes and their antimicrobial resistance profiles circulating in Zambia to guide vaccine development and empirical treatment policies are needed.

## Data Availability

Data underlying the findings are fully available without restriction upon request.

## Acknowledgement

This work is partially funded by the EDCTP2 programme supported by the EU (grant number RIA2018V-2308-ShigaPlexIM). Academic mentorship and data analysis support was received from Professor Dani Cohen and the team from the School of Public Health, Gray Faculty of Medical & Health Sciences, Tel Aviv University, Tel Aviv, Israel.

